# Rising vaccine hesitancy in a future pandemic after COVID-19 and its decision drivers in Japan

**DOI:** 10.64898/2026.01.18.26344375

**Authors:** Junna Kawasaki, Masakaze Hamada, Masaki Machida, Ryo Komorizono, Mie Kobayashi-Ishihara, Naoko Fujita, Takahiro Tabuchi, Yuki Furuse

**Affiliations:** Chiba University; University of Tokyo; Tokyo Medical University; Tohoku University

## Abstract

Vaccine hesitancy emerged as a major challenge during the COVID-19 pandemic and has persisted beyond it, raising concerns about public readiness for vaccination in future pandemics. While extensive research has examined vaccine hesitancy for COVID-19 and routine immunization, it is unknown how the general population would respond to vaccination in a future pandemic, and what conditions might facilitate acceptance. Here, we analyzed data from a nationwide internet-based survey conducted in Japan between December 2024 and January 2025, involving 28,000 participants aged 15–84 years. We assessed intentions to receive vaccination in a hypothetical future pandemic under varying assumptions regarding disease fatality and vaccine immunity durability. Associations between vaccination intention and sociodemographic, psychological, health-related, informational, and infectious disease-related factors were examined. We further explored priority conditions that could increase vaccination intention. We found that only 53.1% of respondents indicated willingness to be vaccinated in a future pandemic with a case fatality rate comparable to COVID-19, representing a marked decline from observed COVID-19 vaccination coverage. Notably, 35.8% of individuals who had vaccinated during the COVID-19 pandemic reported hesitancy toward vaccination in a future pandemic. Higher assumed fatality rates and longer vaccine protection modestly increased willingness, particularly among those with ambivalent attitudes. Vaccination intention was lower among adults aged 20–40 years, women, individuals with lower income or education, and those endorsing misinformation or conspiracy beliefs, while higher infectious disease knowledge, greater fear of COVID-19, and active information seeking were associated with greater willingness. Clustering analysis identified eight distinct groups with heterogeneous priority structures. Free vaccination and clinical trial evidence were universally valued, whereas trust in authorities, domestic vaccine production, and convenience varied substantially across clusters. Public willingness to vaccinate in a future pandemic may be substantially lower than during COVID-19 and is shaped by diverse priorities. Tailored strategies and risk communication approaches that address heterogeneous concerns may be critical for strengthening preparedness for future pandemics.

## Introduction

Vaccination has long been one of the most effective public health measures against infectious diseases, as exemplified by the eradication of smallpox (1,2). Throughout the 20th century, vaccines against poliovirus, measles virus, and many other microbial pathogens were developed to provide direct protection to individuals and to reduce transmission risk through herd immunity. During the COVID-19 pandemic, which began at the end of 2019, vaccines again served as one of the most effective interventions to mitigate clinical and social impact by reducing disease severity.

As of December 2025, more than 700 million infections and over 7 million deaths have been reported worldwide due to COVID-19 (3). Remarkably, after the SARS-CoV-2 genome sequence was released in January 2020, preclinical studies began within two months (4). By December of the same year, the first public vaccinations were initiated in the United Kingdom, followed by the start of vaccination in Japan in February 2021. As of December 2023, approximately 67% of the global population has been vaccinated, with cumulative doses exceeding 13 billion (5). It is estimated that COVID-19 vaccination prevented roughly 14.4 million deaths in the first year of its implementation (6).

The World Health Organization (WHO) identified vaccine hesitancy as one of the ten major threats to global health in 2019 (7). Since the early phase of the COVID-19 pandemic, surveys have examined vaccination intentions and the factors underlying hesitancy to the COVID-19 vaccination (8–11). Global assessments conducted just before and during the initial vaccination rollout reported that approximately 14.3% of individuals were hesitant to receive COVID-19 vaccines (12), with a similar estimate of about 11.3% reported in Japan (13). These reports also indicated that vaccine hesitancy is shaped by concerns about vaccine technology and distrust of governments and pharmaceutical companies, with sociodemographic characteristics contributing to these attitudes. In response, various interventions—such as mandates and financial incentives—have been implemented to increase vaccine uptake, and their effectiveness has been evaluated in multiple studies (14–17).

In addition to traditional platforms such as inactivated and protein subunit vaccines, the COVID-19 pandemic introduced new modalities, including mRNA, viral vector, and replicon-based vaccines (18). Although these modalities raised expectations for improved effectiveness, several studies have reported that they are also associated with higher levels of hesitancy (19,20). In particular, mRNA vaccines have been associated with relatively strong adverse reactions, which may have contributed to increased hesitancy. Previous surveys in Japan indicated that more than 70% of individuals who were hesitant cited concerns about adverse reactions (13). Waning immunity has also been considered a factor promoting hesitancy; for example, the infection-preventive effect of mRNA vaccines declines within several months, and the protection against severe disease diminishes within 6–8 months (21).

The vaccine hesitancy will remain a central challenge in future pandemics. Vaccine hesitancy to even routine immunization has expanded after COVID-19 (22–24), and the reduced vaccine coverage has already caused outbreaks of vaccine-preventable diseases around the world (25,26). The situation has sparked a wide range of discussions about preparedness for the next pandemic in the post-COVID-19 era (27–30). However, quantitative surveys examining attitudes toward vaccination in the context of a potential next pandemic remain scarce. In this study, we surveyed the general population in Japan to assess vaccination intentions, reasons for hesitancy, and strategies to increase vaccine acceptance in a potential next pandemic.

## Methods

### Japan COVID-19 and Society Internet Survey (JACSIS)

An internet survey, JACSIS, was conducted from December 2024 to January 2025. Participants were recruited through a commercial research agency, Rakuten Insight, whose panel comprises approximately 2.2 million individuals from the Japanese population. This survey included 28,000 participants aged between 15 and 84. Details of the recruitment process and sample size calculation were described in our previous protocol paper.

The survey contained 82 head-questions with ∼700 downstream sub-questions; due to branching logic in the questionnaire design, each respondent typically answered approximately 600–700 sub-questions rather than the entire set. Questions in the survey are about demographic information, socioeconomic indicators, health-related factors, psychometric scales, lifestyle behaviors, social measures, and COVID-19- and other infectious diseases-related measures (e.g., infection history, preventive behaviors, vaccination status, vaccine attitudes, fear of COVID-19, knowledge about infectious diseases). All the questions in the survey are available on our website (https://jacsis-study.jp/dug/index.html).

### Outcome

Intention to be vaccinated in the next pandemic was asked with four answering options of “Definitely will be vaccinated,” “Probably will be vaccinated,” “Probably won’t be vaccinated,” and “Definitely won’t be vaccinated.” The questions were asked under different scenarios for the disease’s fatality and the durability of vaccination immunity. The details of the question are provided in Supplementary text S1.

### Variables for vaccination intention

Association with vaccination intention in the next pandemic was investigated for demographic information (age and sex), socioeconomic status (income and educational level), contact patters with others (job, living status, marital status, and contact frequency), healthcare and medical-related factors (consultation with a regular doctor, the presence of comorbidity, COVID-19 infection history, and COVID-19 vaccination adverse events), disposition and personality (levels of anxiety and happiness, the fear of COVID-19, and communion and agency orientations; the criteria to define communion and agency orientations are explained in the Supplementary text S2), infectious diseases-related perceptions (knowledge of infectious diseases, beliefs in misinformation and conspiracy statements, and the expectation about the timing of the next pandemic; details are described in Supplementary text S3), and information sources for COVID-19.

The associations were tested for vaccine intention, excluding individuals who could not be vaccinated for medical reasons, such as allergies. That was done because we could not tell whether the response about their vaccination intention was based on psychological or physical acceptance. “Definitely will be vaccinated” and “Probably will be vaccinated” were grouped as “Will”, and “Probably won’t be vaccinated” and “Definitely won’t be vaccinated” were merged as “Won’t.” Statistical adjustment was conducted for age, sex, income, and educational level using binomial regression in R (version 4.3.2). Covariates were selected a priori based on substantive knowledge. The 95% confidence intervals were calculated, and P-values < 0.05 were considered statistically significant.

### Facilitating factors for vaccination

The survey also investigated factors or conditions that can enhance vaccination intention. We asked whether 20 items could enhance willingness (Supplementary text S4). Response options are “1. My intention to receive the vaccine would increase very strongly,” “2. My intention to receive the vaccine would increase somewhat,” “3. This would not have much influence on my decision about whether to receive the vaccine,” “4. My intention not to receive the vaccine would increase somewhat,” “5. My intention not to receive the vaccine would increase very strongly.”

To quantitatively analyze priorities of the 20 factors among individuals, the median score for the 20 questions was calculated for each participant. Then, the scores for the 20 questions were converted to the difference from the median value. After the conversion, unsupervised clustering was performed, computing a dissimilarity matrix with Gower’s distance. Then, partitioning around medoids clustering was applied across a range of cluster numbers (k = 2–20) in order to determine the optimal number of clusters. For each k, the average silhouette width was calculated to assess cluster separation and cohesion. The number of clusters that maximized the average silhouette width was selected as the optimal solution. The computation was performed using the cluster and gower packages in R.

### Ethical statements

The JACSIS study received approval from the Institutional Review Board of the Osaka International Cancer Institute (approval number: 20084), from the Ethics Committee of Tohoku University Graduate School of Medicine (2024-1-517), and from the Research Ethics Committee of the Graduate School of Medicine and Faculty of Medicine at the University of Tokyo (2020336NI).

All participants provided informed consent before taking part in the survey; the consent process was conducted online. For participants aged 15–17 years, informed consent was obtained directly from them without parental consent in accordance with the Ethical Guidelines for Medical and Biological Research Involving Human Subjects implemented by Japan’s Ministry of Education, Culture, Sports, Science and Technology and Japan’s Ministry of Health, Labour and Welfare.

## Results

### Potential for stronger vaccine hesitancy in a future pandemic relative to COVID-19

Among the 28,000 participants, we excluded satisficing or inconsistent responses, yielding 25,082 responses for our analysis in the study (a valid response rate of 89.6%). Participants were aged 15 to 84 years. Of these, 19,027 individuals had received at least one dose of a COVID-19 vaccine, accounting for 75.9% of the total (19,027/25,082), a proportion consistent with the national vaccination coverage reported in 2022 (80.4%) (31,32). For subsequent analyses, we further excluded 505 respondents who did not receive the COVID-19 vaccination due to medical reasons, such as allergies, resulting in a final analytical sample of 24,577 individuals.

We first assessed intentions to receive vaccination in a hypothetical future pandemic. Figure 1A shows vaccination intention assuming that the case fatality rate of the next pandemic would be comparable to that of COVID-19 (approximately 0.1–2%). Based on two questions—whether respondents had received COVID-19 vaccination and whether they would choose to be vaccinated in the next pandemic—we categorized individuals into four groups: “Did–Will,” “Didn’t–Will,” “Did–Won’t,” and “Didn’t–Won’t.”

**Figure 1.**
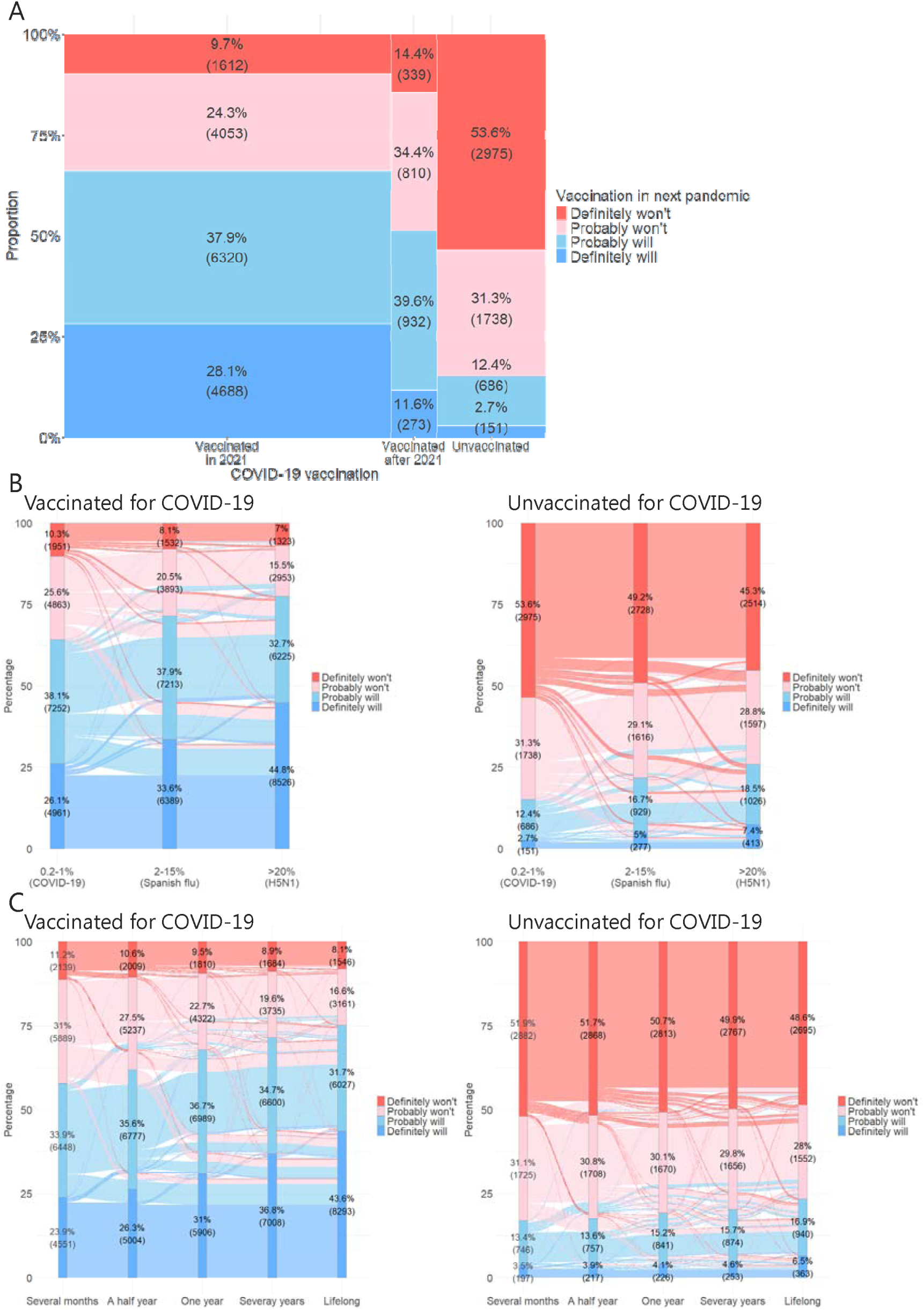
Vaccination intention in a future pandemic. The proportions and numbers of respondents for each vaccination-intention option in a future pandemic are shown. A) An assumed pathogenicity of the hypothetical pandemic is equivalent to COVID-19. The results are stratified by COVID-19 vaccination status and the timing of the first dose. B) Hypothetical scenarios are changed for the pathogenicity of the pandemic, and C) the durability of vaccination immunity.

In this survey, 53.1% of respondents (13,050/24,577) reported that they would be willing to receive vaccination in a future pandemic. Among those vaccinated during the COVID-19 pandemic, 35.8% (6,814/19,027) indicated they would hesitate to receive vaccination in a future pandemic (“Did–Won’t” group). This tendency was even stronger among individuals who were unvaccinated during the COVID-19 pandemic; 84.9% (4,713/5,550) reported that they would not intend to be vaccinated in the next pandemic (“Didn’t–Won’t” group).

The degree of hesitancy varied depending on the assumed pathogenicity and the duration of vaccine protection (Figure 1BC). Particularly, individuals in the “Probably won’t” group showed substantial shifts toward willingness to be vaccinated. As the assumed fatality rate increased, the proportion of “Probably won’t” declined from 25.6% to 15.5% among those vaccinated against COVID-19 and from 31.3% to 28.8% among COVID-19-unvaccinated individuals. Similarly, with a longer assumed duration of protection from vaccination, the “Probably won’t’ proportion decreased from 31.0% to 16.6% among COVID-19-vaccinated individuals and from 31.1% to 28.0% among the unvaccinated. Even some individuals in the “Definitely won’t” group shifted toward vaccine acceptance, particularly when faced with high fatality of a pandemic or long-term vaccine effectiveness, although the magnitude was limited.

### Individual characteristics associated with vaccine hesitancy

We next examined the associations between individual characteristics and vaccination intention for the next pandemic using a binomial logistic regression analysis adjusted for age, sex, income, and educational level. We first focused on sociodemographic indicators (Figure 2A and Supplementary table S1). Vaccination intention was lower among individuals in their 20s to 40s, and women also showed a lower odds of intending to be vaccinated in a future pandemic. Lower income and lower educational attainment were similarly associated with reduced vaccination intention.

**Figure 2.**
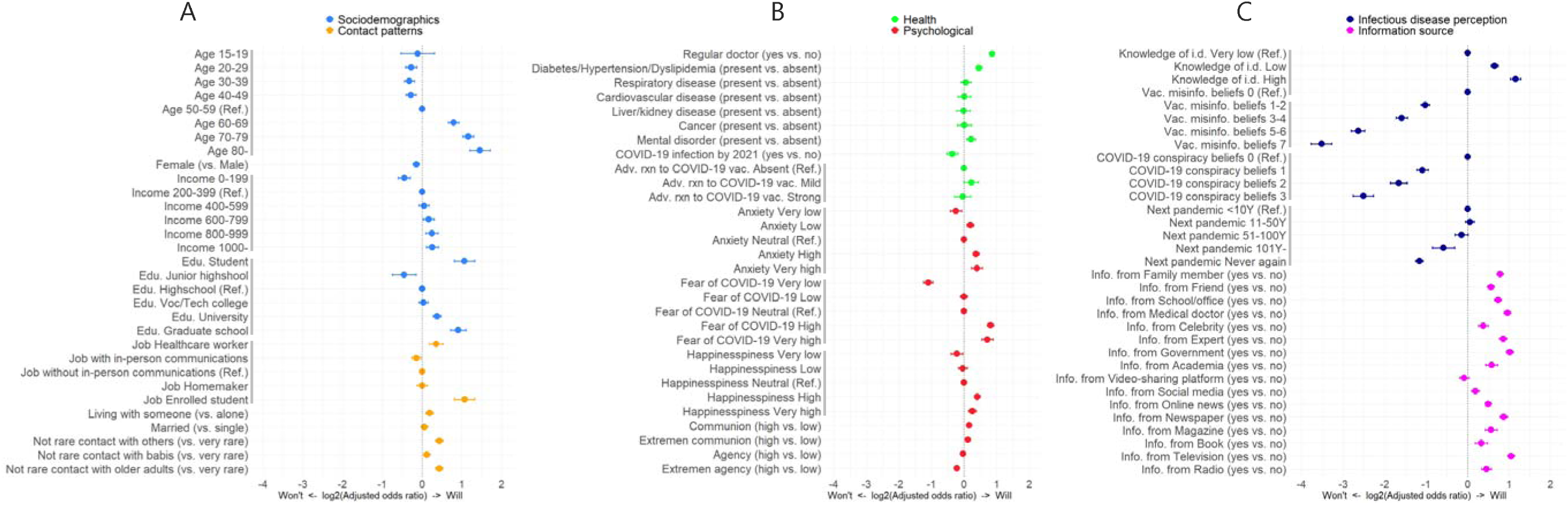
Association of variables with vaccination intention in a future pandemic. Associations of variables for the vaccination intention are shown by odds ratios adjusted for age, sex, income, and education. Horizontal lines indicate 95% confidence intervals. Variables are categorized in sociodemographics, contact patterns (A), healthcare/medical-related factors, psychological factors (B), infectious disease-related perception, and information sources (C). Income is expressed in x10,000 Japanese yen. Edu., Educational level; Adv. rxn to COVID-19 vac., Adverse reaction after COVID-19 vaccination; i.d., infectious diseases; Info., Information source.

We also evaluated whether interpersonal contact influenced vaccination intention. Regarding their job, healthcare workers and enrolled students demonstrated a higher willingness to be vaccinated in the next pandemic. Individuals who reported non-rare contact with others were more likely to intend to receive vaccination, with particularly high willingness observed among those who frequently interacted with older adults.

We further examined the relationship between vaccination intention and healthcare/medical factors (Figure 2B). Individuals who see a regular physician were more willing to be vaccinated. Among the listed comorbidities, only cardiometabolic diseases, including diabetes, hypertension, and dyslipidemia, and mental disorders were significantly associated with vaccination intention, whereas other conditions did not show significant effects. Interestingly, neither the presence nor the severity of adverse reactions by COVID-19 vaccination influenced future vaccination intention in this survey.

Analysis of psychological factors showed that individuals who reported lower levels of general anxiety tended to exhibit stronger vaccine hesitancy. A similar pattern was observed for fear of COVID-19: those who experienced little fear were more likely to be hesitant. Examination of communion/agency orientation personality indicated that individuals with higher communion (including extreme communion) were more willing to be vaccinated, whereas those with extreme agency were more likely to avoid vaccination.

We assessed how perceptions of infectious diseases influenced vaccination intention in the next pandemic (Figure 2C). Participants who reported greater knowledge of infectious diseases and vaccines showed greater willingness to vaccinate, whereas those who endorsed vaccine-related misinformation or COVID-19 conspiracy beliefs exhibited stronger hesitancy. Additionally, individuals who believed that another pandemic would not occur within the next 100 years were more likely to be hesitant.

Across the 16 information sources explored, respondents who reported actively seeking COVID-19-related information generally showed higher vaccination intention, except for video-sharing platforms. No information source was associated with increased vaccine hesitancy in this survey. Among the information sources, the government, healthcare professionals, medical experts, television, and newspapers were particularly effective in promoting vaccine acceptance.

### Subgroup-specific determinants of vaccination intention

Next, we examined vaccination-related conditions that can affect willingness to be vaccinated across eight major domains (Figure 3). As items related to medical and scientific evidence, we assessed types of clinical trial data (A). Recommendations are divided into recommendations from public health and medical institutions (B) and recommendations from peers (C). As vaccine development-related conditions, we evaluated the development or manufacturing country (D) and vaccine modality (E). Regarding access and convenience, we investigated administrative simplicity (F) and accessibility in terms of time and location (G). Finally, as an economic factor, we examined the cost of vaccination (H). We checked the proportion of individuals who answered that the indicated condition can increase vaccination intention. Overall, most individuals in the “Did–Will” group perceived most conditions as factors that would increase vaccination intention, whereas in the “Didn’t–Won’t” group, such responses were limited to approximately 15% (Figure 3A).

**Figure 3.**
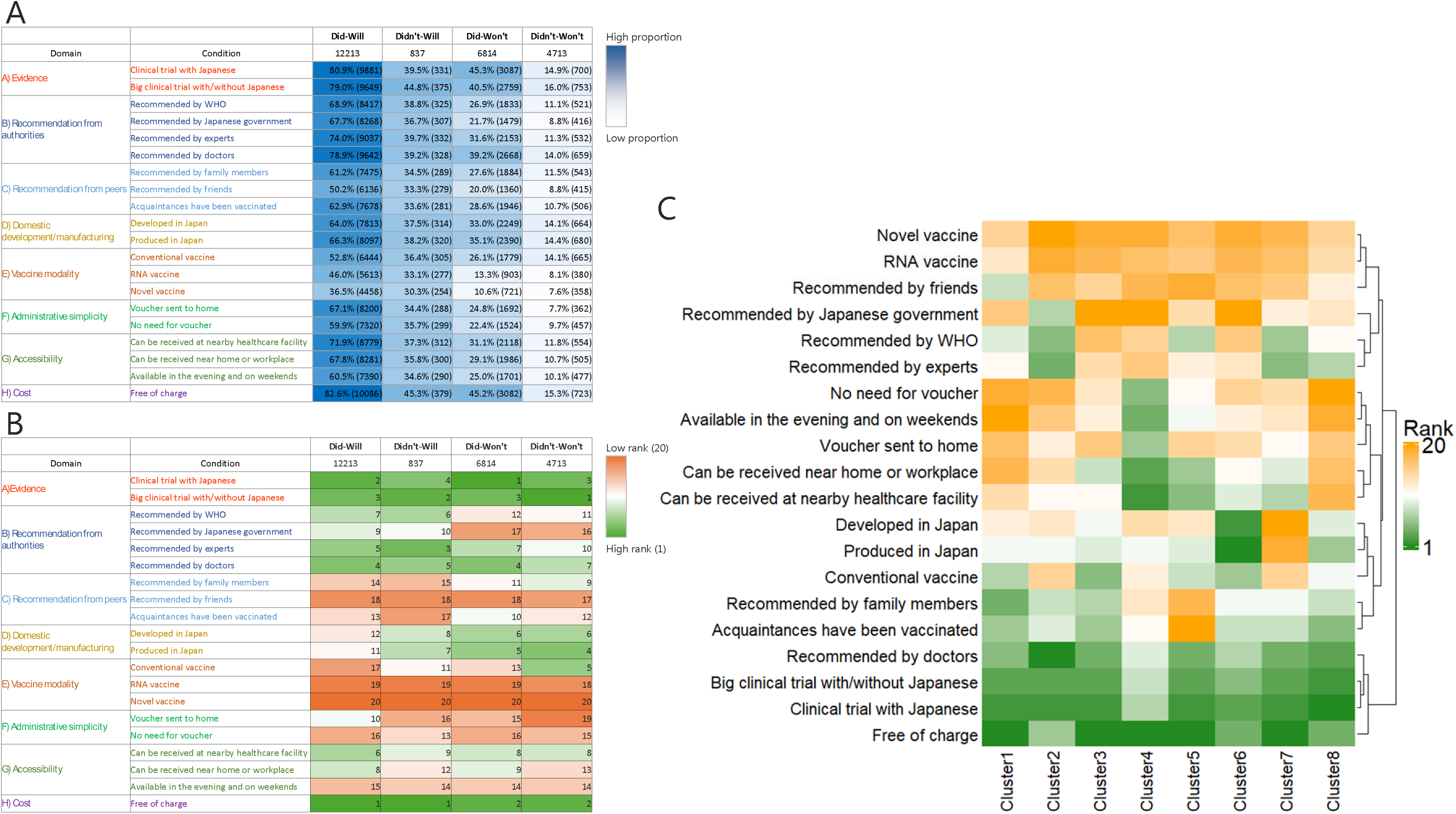
Conditions that affect vaccination intention in a future pandemic. A) The proportions and numbers of respondents who answered that an indicated condition can enhance the vaccination intention in a future pandemic are shown for four vaccination intention groups. B) Priority ranking of the twenty conditions is shown. C) The result of unsupervised clustering of individuals based on the prioritization ranking of each condition is shown. Twenty conditions are sorted by hierarchical similarity, and eight clusters are sorted by the size of the individuals.

Interestingly, when examining which conditions were more valued for increasing vaccination intention within each group, similar and distinct patterns emerged between the “Will’’ and “Won’t’’ groups (Figure 3B). The most influential factors included “free of charge” and the items related to medical and scientific evidence for all groups. In contrast, recommendations from the WHO and the Japanese government were particularly valued in the “Will’’ groups but were given little weight in the “Won’t’’ groups. Among the “Won’t” groups, the vaccine being developed or manufactured domestically, i.e., in Japan, appeared to exert a relatively greater influence on vaccination intention, whereas the “Did–Will’’ group placed little emphasis on whether the vaccine was domestically produced, highlighting clear differences in priority structures among the groups.

### Priority conditions for vaccination intention differ across individuals

Clustering analysis of priority conditions for vaccination in a future pandemic revealed that participants could be classified into eight clusters (Figure 3C and Table 1). Across all clusters, free vaccination, the evidence from clinical trial data, and physician recommendations were consistently valued, whereas newer vaccine modalities, such as RNA vaccines, did not substantially increase vaccination intention. This pattern was also evident in Figure 3B. In contrast, the importance of convenience varied considerably among clusters. Overall, conditions related to time and location accessibility were prioritized more frequently than those related to administrative simplicity.

**Table 1.** Summary of clustering on prioritized conditions for vaccination intention.

| Cluster ID | 1 | 2 | 3 | 4 | 5 | 6 | 7 | 8 |
| --- | --- | --- | --- | --- | --- | --- | --- | --- |
| Summary of prioritized conditions | (More influenced by others)<br>Little care about admin. simplicity or accessibility<br>(Less concern in novel technologies) | Much trust in authority<br>Little care about admin. simplicity or accessibility<br>(Less care about price) | Little trust in authority<br>Little care about admin. simplicity | Much care about admin. simplicity and accessibility<br>Little trust in authority<br>(Less care about scientific evidence) | Much care about accessibility<br>(More trust in authority)<br>Little influenced by others | Much care about Japan-made<br>Little trust in authority | Much trust in authority<br>Little care about Japan-made | Little care about admin. simplicity or accessibility<br>(More trust in authority)<br>(More care about Japan-made)<br>(Less concern in novel technologies) |
| Numbers of participants | 11,620 | 2,385 | 1,917 | 1,917 | 1,828 | 1,801 | 1,761 | 1,348 |
| Did-Will | 39.2% (4,553) | 61.7% (1,472) | 55.2% (1,058) | 51.9% (995) | 66.6% (1,217) | 44.9% (808) | 74.0% (1,304) | 59.8% (806) |
| Didn't-Will | 4.0% (466) | 2.7% (64) | 2.8% (53) | 4.0% (77) | 2.6% (47) | 2.7% (49) | 1.9% (34) | 3.5% (47) |
| Did-Won't | 29.3% (3,399) | 25.8% (616) | 28.6% (549) | 27.9% (534) | 22.3% (408) | 38.1% (686) | 16.5% (290) | 24.6% (332) |
| Didn't-Won't | 27.6% (3,202) | 9.8% (233) | 13.4% (257) | 16.2% (311) | 8.5% (156) | 14.3% (258) | 7.6% (133) | 12.1% (163) |
| Profiling | Low education,<br>Less contact with others,<br>Little info. gathering,<br>Low fear of COVID-19,<br>Less comorbidity,<br>Less knowledge of I.D.<br>More misinfo. Beliefs | More unemployed,<br>Less misinfo. beliefs | More female,<br>More misinfo. beliefs,<br>More info. from online | Less older adults,<br>More single,<br>Low happiness,<br>More misinfo. beliefs | Good knowledge of i.d.,<br>More info. from authorities, | More misinfo. beliefs | High education,<br>Good knowledge of i.d.,<br>Little misinfo. beliefs<br>More info. from authorities | More older adults, High happiness,<br>More comorbidity |

Admin., administrative: i.d., infectious disease; info., information; misinfo., misinformation

Profiling of each cluster revealed that Cluster 1 comprised nearly half of all participants (47.28%, 11,620/24,577) and showed the highest proportion of individuals in the “Won’t” group (56.8%, 6,601/11,620) (Table 1 and Supplementary table S2). This cluster placed relatively low importance on convenience-related conditions but assigned higher priority to recommendations from family and friends. Clusters 2, 5, and 7 contained higher proportions of individuals in the “Did–Will’’ group. Clusters 2 and 7 tended to value guidance from professional or expert institutions, and Clusters 5 and 7 showed higher scores on infectious disease knowledge and included more highly educated respondents. Despite their broadly similar profiles in Clusters 5 and 7, these two clusters differed in the priority assigned to certain conditions. Cluster 5 does not prioritize recommendations from family and acquaintances, while Cluster 7 possesses no particular preference in domestically produced vaccines or traditional vaccine modality. Cluster 8 resembled Cluster 1 in assigning low priority to convenience but differed in its relatively stronger preference for domestically produced vaccines; this cluster included a larger proportion of older adults.

Clusters 3, 4, and 6 placed less importance on recommendations from authorities and had higher proportions of individuals in the “Won’t’’ groups, followed by Cluster 1. Traditional vaccine modality is relatively prioritized in Clusters 3 and 6. Cluster 6 was further characterized by very strong preferences for domestically produced vaccines, similar to Cluster 8. Cluster 4 was characterized by a high priority for convenience-related conditions. This cluster contained a relatively large proportion of younger individuals, suggesting that convenience plays a particularly important role for certain segments of younger respondents.

## Discussion

In this study, we conducted a large-scale survey in Japan to assess vaccination intentions in a future pandemic. Although a recent study using data from 1 million people in the United Kingdom reported the prevalence and relationship between initial attitudes toward COVID-19 vaccination and subsequent vaccine uptake, it investigated the mechanisms of vaccine hesitancy and effective strategies within the context of COVID-19 (33). Our survey revealed that only 52.7% of the participants indicated willingness to receive vaccination in a future pandemic with an assumed case fatality rate comparable to that of COVID-19. Given that COVID-19 vaccination coverage in Japan reached >80% in 2022, this decline in vaccination intention poses a significant challenge to preparedness for the next pandemic. Notably, 35.8% of individuals who had been vaccinated during the COVID-19 pandemic shifted to the hesitant group when asked about a hypothetical future pandemic. Understanding these patterns of vaccine hesitancy and establishing systems that enable appropriate vaccine delivery will be essential components of future pandemic preparedness.

To investigate the relationship between individual characteristics and vaccination intention, we examined a range of demographic, social, psychological, and health-related factors. Consistent with previous research, various attributes—including age, sex, and frequency of interpersonal contact—were associated with willingness to vaccinate (11). An interesting finding of this study is that neither the presence nor the severity of adverse reactions was negatively associated with vaccination intention. Prior studies have similarly reported that mild adverse effects do not reduce willingness to vaccinate, and some individuals even interpret such reactions as evidence of a good immune reaction to the vaccine (34–36).

Furthermore, we found that the delivery of information must play a substantial role. As highlighted in earlier studies and ours, belief in vaccine-related misinformation or COVID-19 conspiracy theories was strongly associated with vaccine hesitancy (11,37). Our survey unveiled that particular information sources are more effective at enhancing vaccine acceptance (Figure 2C), although reliance on these sources differs substantially among individuals (Figure 3C). These findings may underscore the importance of disseminating accurate information regardless of the communication platform.

As such, one of the key contributions of this study is that individuals who showed reluctance to vaccination may still have a chance to change their minds. We found that 15.1% of the even COVID-19 unvaccinated people showed the willingness to be vaccinated in a future pandemic (Figure 1A), and hesitancy levels shifted depending on the assumed case-fatality rate for the next pandemic and the expected duration of vaccine protection (Figure 1BC). The change from hesitancy to acceptance is large especially among those who got vaccinated for COVID-19, suggesting their volatile, teetering attitude. A previous study identified differences in the characteristics of hesitant individuals between those who later accepted vaccination and those who remained more resistant (33). Furthermore, the present study identified conditions that are prioritized differently by already-vaccine-accepting and currently-vaccine-hesitant groups.

Free vaccination and the presence of clinical trial data were consistently prioritized across all groups (Figure 3BC). In contrast, recommendations from authoritative organizations such as the WHO, the development of domestically produced vaccines, and factors related to convenience and accessibility exhibited substantial individual variability in their influence on vaccination intention. Understanding these group-level differences in prioritized vaccination conditions may provide a useful approach for addressing vaccine hesitancy. For example, administrative simplicity and easy access can promote vaccination in younger generations, while domestic development can appeal to older individuals. Those who recommend vaccination affect differently among individuals, suggesting a polyhedral approach is warranted.

This study has several limitations. First, the survey was conducted in Japan during a specific period, and further investigation is needed to determine the generalizability of the findings. To address this limitation, continued longitudinal surveys and expansion to multi-country settings will be needed. As participants were recruited via an online platform, selection bias cannot be ruled out. Furthermore, although this study showed that only approximately 50% of respondents expressed willingness to be vaccinated in a future pandemic, this proportion should not be interpreted as a precise estimate of future vaccination coverage. Indeed, previous research has noted discrepancies between early vaccination intentions and actual uptake during the COVID-19 vaccine rollout (38,39). Finally, a comprehensive understanding of vaccine hesitancy will require the integration of qualitative approaches alongside quantitative analyses.

Our survey, which assumed a future pandemic, revealed a substantial increase in the proportion of individuals exhibiting vaccine hesitancy. These hesitant groups appear to differ from vaccine-accepting groups not only in their individual characteristics but also in the conditions they prioritize when considering vaccination. Our findings can help develop more individualized strategies for vaccination delivery. At the same time, the present study underscores the need to develop effective risk communication strategies tailored to target populations. By identifying risk factors related to vaccine hesitancy during non-pandemic periods, this study contributes to preparedness for future pandemics.

## Data Availability

All data produced in the present study are available upon reasonable request to the authors.

## Acknowledgments

The Japan Agency for Medical Research and Development (JP223fa627001, JP223fa627004, 23wm0125011)

The Japan Society for the Promotion of Science (JP21H04856, JP23K09693, JP25H01079, 120248602)

The Ministry of Health, Labour and Welfare of Japan (22JA1005, 22FA1001, 22FA1010, 23EA1001, 23FA1004, 24FA1020, 24LA1002)

The Public Foundation of Vaccination Research Center (NA)

## Supplementary materials

**Supplementary table S1.**
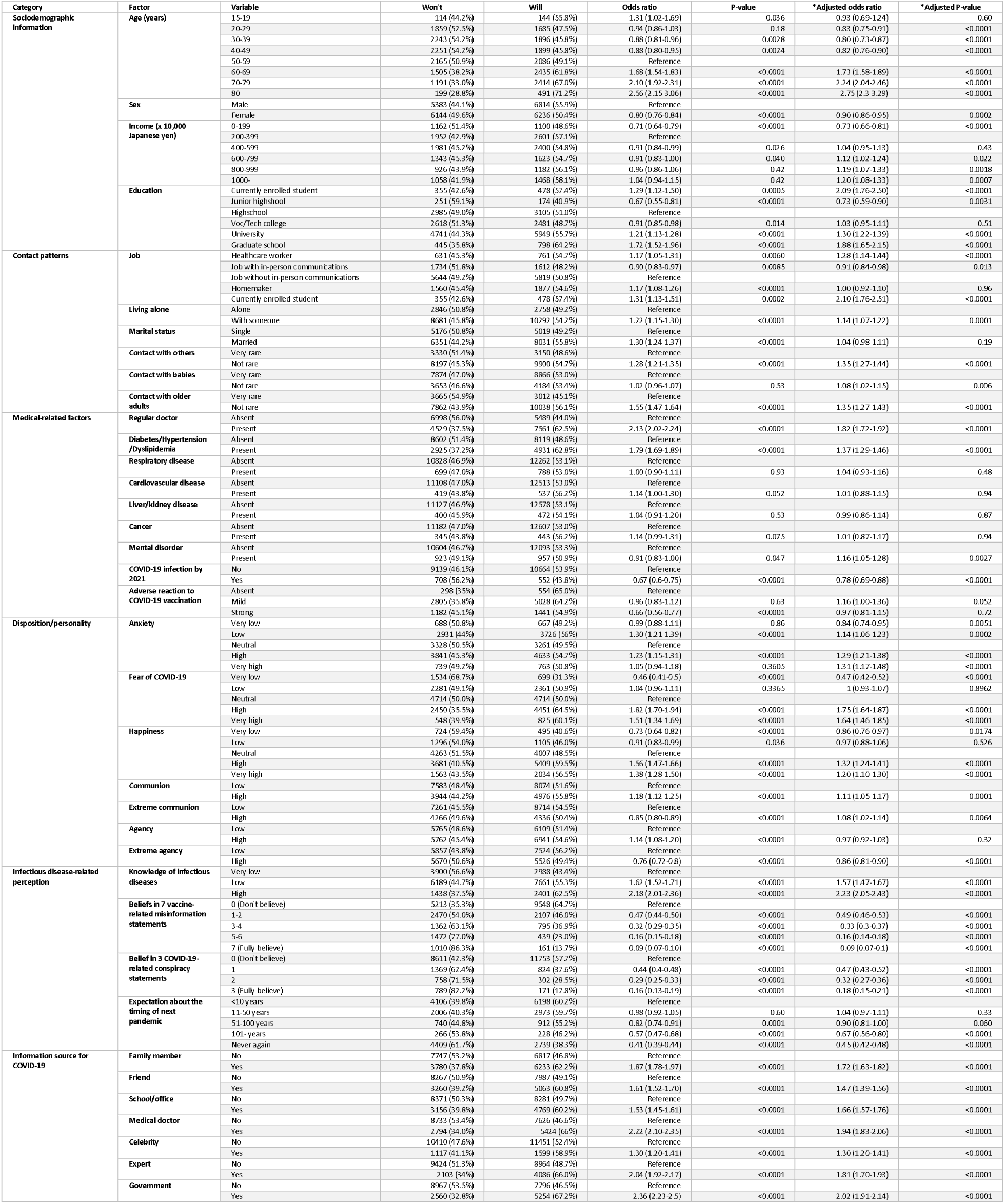
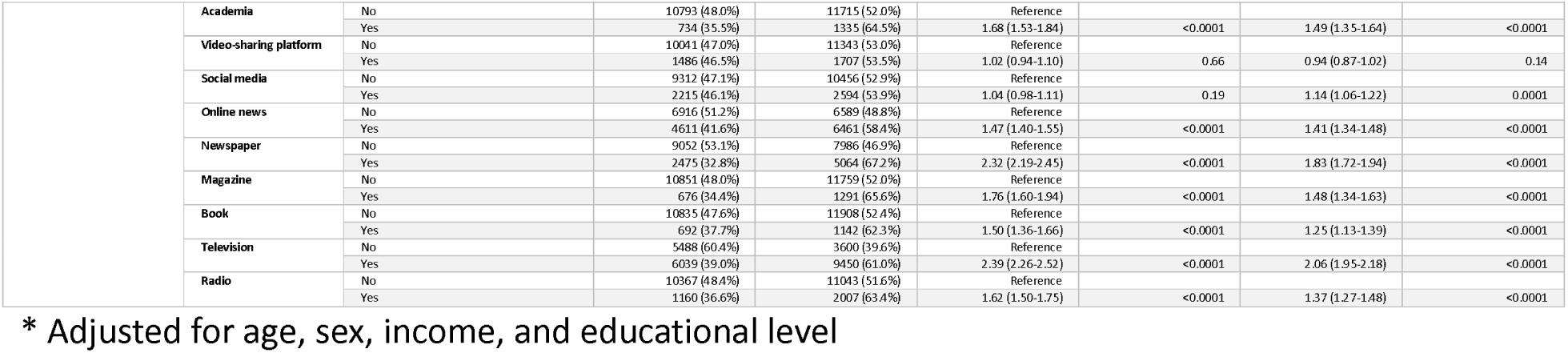
Association of variables with vaccination intention for the next pandemic.

**Supplementary table S2.**
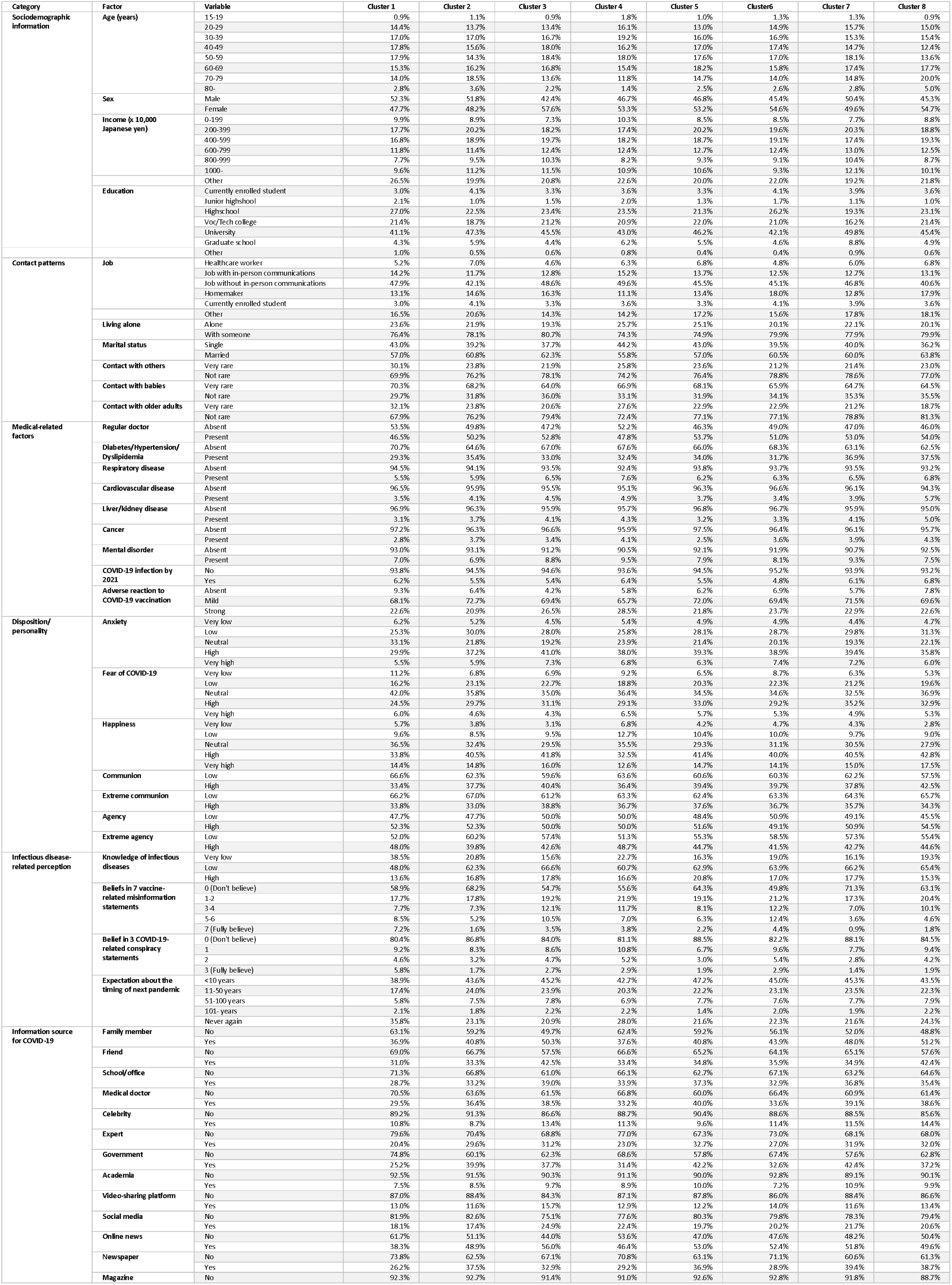
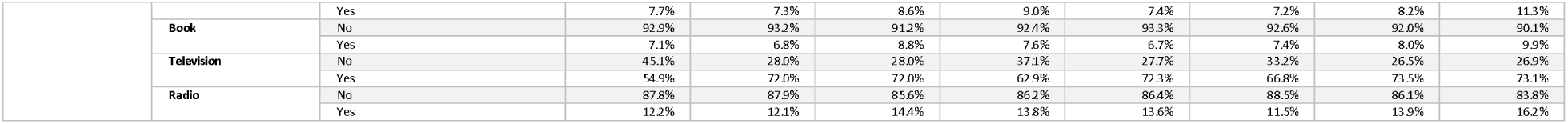
Profile of clusters based on the priority of facilitating factors.

### Supplementary text S1 Questions about vaccination intention for the next pandemic

Please answer the following questions assuming that the next pandemic occurs. It is assumed that a vaccine capable of reducing the risk of death from this pandemic by more than 50% has been developed, and that it has been approved by the Japanese government, the WHO (World Health Organization), and other relevant authorities. Under the following conditions regarding the case fatality rate of the pathogen causing the pandemic (i.e., the proportion of infected individuals who die) and the duration of vaccine effectiveness, would you like to receive the vaccine?

- In the case of a pandemic with a case fatality rate comparable to that of COVID-19 (approximately 0.1–2%)
- In the case of a pandemic with a case fatality rate of approximately 2–15% (comparable to the Spanish influenza pandemic of 1918 or the SARS outbreak that spread overseas in 2003)
- In the case of a pandemic with a case fatality rate of 20% or higher (comparable to highly pathogenic avian influenza or Ebola virus disease)
- In the case of a pandemic with a case fatality rate of 2%, and the vaccine effectiveness is maintained for 2–3 months
- In the case of a pandemic with a case fatality rate of 2%, and the vaccine effectiveness is maintained for about six months
- In the case of a pandemic with a case fatality rate of 2%, and the vaccine effectiveness is maintained for about one year
- In the case of a pandemic with a case fatality rate of 2%, and the vaccine effectiveness is maintained for several years
- In the case of a pandemic with a case fatality rate of 2%, and the vaccine effectiveness lasts for a lifetime

### Supplementary text S2 Questions about variables for communion/agency orientation

The questions and scoring system were proposed and validated for Japanese individuals in published studies:

https://www.jstage.jst.go.jp/article/jjpsy1926/75/5/75_5_420/_article

https://www.jstage.jst.go.jp/article/pacjpa/71/0/71_3PM143/_article

https://shoin.repo.nii.ac.jp/record/1578/files/K49-2Dohirakawamizusawa.pdf

For each question, participants were asked to select one of “1. Strongly disagree,” “2. Disagree,” “3. Agree,” “4. Strongly agree.”

Communion:

- I express words of gratitude verbally.
- I am able to think from the other person’s perspective.
- I am good at complimenting others.
- I am able to apologize honestly.
- I am able to cooperate with others.
- I treat others with compassion and consideration.

Scores of 19 or higher were considered to indicate communion orientation.

Extreme communion:

- I am not good at expressing my opinions in front of others.
- I worry too much about other people.
- I tend to think about relying on others right away.
- I think too much about people around me and become unable to take action.
- I am easily hurt by what others say.
- I tend to overinterpret what others say.

Scores of 17 or higher were regarded as extreme communion orientation.

Agency:

- I am proactive and take initiative in my activities.
- I assert my own opinions.
- I am confident in myself.
- I do not give up even when I face difficulties.
- Once I make up my mind, I act immediately.
- I have strong willpower and firm beliefs.

Scores of 16 or higher were considered indicative of agency orientation.

Extreme agency:

- I cannot tolerate incompetent people.
- I make others do what I want.
- I take an aggressive attitude toward others.
- I do not listen to the other person’s point of view.
- I am unable to accept opinions that differ from my own.
- I cannot forgive other people’s mistakes.

Scores of 13 or higher were regarded as extreme agency orientation.

### Supplementary text S3 Questions about variables related to infectious diseases

Adverse reaction after COVID-19 vaccination:

Did any of the following occur within 1–3 days after receiving the COVID-19 vaccine?

- Pain at the injection site
- Fatigue or general malaise
- Headache
- Muscle pain or joint pain
- Chills
- Fever
- Swelling at the injection site
- Nausea or vomiting
- Other symptoms

Response options are “1. Did not occur,” “2. Occurred, but I did not visit a hospital and did not miss school or work,” “3. Occurred, so I visited a hospital and missed school or work,” “4. Occurred, so I missed school or work but did not visit a hospital,” and “5. Occurred, so I visited a hospital but did not miss school or work.” Respondents who chose option 1 for all questions were regarded as having an “absent” adverse reaction, those who selected option 3 or 4 for at least one question were considered as having a “strong” adverse reaction, and the others were classified as having a “mild” adverse reaction.

Knowledge of infectious diseases:

- Most cases of cervical cancer are caused by HPV (human papillomavirus).
- HPV (human papillomavirus) is generally transmitted through sexual activity.
- Approximately one in four people who develop cervical cancer die from the disease.
- HPV infection can also cause cancer in men.
- The Ministry of Health, Labour and Welfare actively recommends HPV vaccination.
- Girls and women aged 17–27 who did not receive the HPV vaccine because active recommendations were withheld can receive the vaccine free of charge.
- A new vaccine (Gardasil 9) was introduced into the routine immunization program starting in fiscal year 2023.
- If health damage occurs due to adverse reactions to vaccination, and a causal relationship between the vaccination and the health damage is recognized, there is a public compensation system that covers the medical expenses incurred and other related costs such as hospitalization and outpatient care.
- If one wishes to receive compensation for health damage caused by adverse reactions to vaccination, one should consult the local municipal government regarding the procedures.
- Men aged 45–62 in the current fiscal year did not have the opportunity to receive rubella vaccination during childhood.
- The government recommends rubella vaccination (routine immunization) for men aged 45–62.
- In the routine rubella vaccination program for adult men, a free blood test (antibody test) can be performed in advance.
- If a pregnant woman becomes infected with rubella, there is a possibility that her baby will be born with congenital rubella syndrome. (Congenital rubella syndrome refers to congenital disorders such as hearing impairment, congenital heart defects, visual impairment, and delays in mental or physical development.)
- Respiratory syncytial virus (RS virus) is a virus that causes respiratory infections such as bronchitis and pneumonia.
- Almost all children are infected with RS virus at least once by the age of two.
- Among children under two years of age who visit a hospital due to RS virus infection, approximately one in four requires hospitalization.
- By receiving the RS virus vaccine during pregnancy, it is possible to prevent severe RS virus infection in the child who will be born.
- Pregnant women are eligible to receive the RS virus vaccine.
- Older adults and people with underlying medical conditions are at higher risk of developing severe RS virus infection.
- Older adults are eligible to receive the RS virus vaccine.

Response options include “I knew” and “I did not know.” Individuals who knew 0, 1–10, 11–20 statements were regarded having “very low,” “low,” and “high” knowledge about infectious diseases.

Seven vaccine-related misinformation:

- Data on vaccine safety are often fabricated.
- Childhood vaccination is harmful, and this fact is being concealed.
- Pharmaceutical companies are hiding the dangers of vaccines.
- People are being deceived about the effectiveness of vaccines.
- Data on vaccine effectiveness are often fabricated.
- People are being deceived about vaccine safety.
- The government is trying to hide a link between vaccines and autism.

Response options include “Strongly disagree,” “Disagree,” “Neither agree nor disagree,” “Agree,” and “Strongly agree.” Respondents who endorsed (selected “Agree” or “Strongly agree”) 0, 1–2, 3–4, 5–6, and 7 statements were considered to have “zero,” “low,” “middle,” “high,” and “very high” misinformation beliefs, respectively.

Three COVID-19-related conspiracy statements:

- Large pharmaceutical companies created COVID-19 in order to profit from vaccines.
- COVID-19 was created to force all people to get vaccinated.
- They are trying to carry out mass sterilization using this vaccine.

Response options include “Strongly disagree,” “Disagree,” “Neither agree nor disagree,” “Agree,” and “Strongly agree.” Respondents who endorsed (selected “Agree” or “Strongly agree”) 0, 1, 2, and 3 statements were considered to have “zero,” “low,” “middle,” and “high” conspiracy theory beliefs, respectively.

Expectation about the next pandemic:

- When do you think the next pandemic (a global outbreak of an infectious disease) like COVID-19 is most likely to occur?

The available response choices included: “within 1–2 years,” “in 3–5 years,” “in 6–10 years,” “in 11–50 years,” “in 51–100 years,” “more than 100 years from now,” or “it will never occur.

### Supplementary text S4 Twenty possible conditions facilitating vaccination intention

- The safety and effectiveness have been confirmed through clinical studies (clinical trials) that included Japanese participants.
- The safety and effectiveness have been confirmed through large-scale clinical studies, regardless of whether Japanese participants were included.
- The WHO (World Health Organization) recommends vaccination.
- The Japanese government recommends vaccination.
- Many experts recommend vaccination.
- Many physicians and healthcare professionals recommend vaccination.
- My family recommends vaccination.
- My friends or colleagues recommend vaccination.
- My family members or acquaintances have been vaccinated.
- The vaccine was developed in Japan.
- The vaccine is manufactured in Japan.
- The vaccine is produced using conventional methods, such as protein-based vaccines, inactivated vaccines, or live attenuated vaccines.
- The vaccine is an RNA vaccine, a relatively new technology that was widely used for COVID-19 vaccines.
- The vaccine is based on a new technology that has not previously been put into practical use.
- A vaccination voucher is sent to my home.
- No vaccination voucher is required, and vaccination can be received by going directly to the vaccination site.
- Vaccination can be received at my regular healthcare provider or at a nearby medical facility.
- Vaccination can be received near my home or workplace, or at my workplace.
- Vaccination is available at night or on holidays.
- Vaccination is free of charge.

## Notes

### Competing Interest Statement

The authors have declared no competing interest.

### Funding Statement

This study was funded by the Japan Agency for Medical Research and Development (JP223fa627001, JP223fa627004, 23wm0125011), the Japan Society for the Promotion of Science (JP21H04856, JP23K09693, JP25H01079, 120248602), the Ministry of Health, Labour and Welfare of Japan (22JA1005, 22FA1001, 22FA1010, 23EA1001, 23FA1004, 24FA1020, 24LA1002), and the Public Foundation of Vaccination Research Center (grant number not available).

### Author Declarations

Ethics committee/IRB of the Osaka International Cancer Institute, Tohoku University Graduate School of Medicine, and the Graduate School of Medicine and Faculty of Medicine at the University of Tokyo gave ethical approval for this work.

## References

1. World Health Organization. WHA33.3 - Declaration of global eradication of smallpox [Internet]. World Health Organization; 1980 [cited 2026 Jan 10]. Available from: https://www.who.int/publications/i/item/WHA33-3

2. World Health Organization. A Brief History of Vaccination [Internet]. [cited 2026 Jan 10]. Available from: https://www.who.int/news-room/spotlight/history-of-vaccination/a-brief-history-of-vaccination

3. World Health Organization. WHO COVID-19 dashboard [Internet]. datadot. [cited 2025 June 5]. Available from: https://data.who.int/dashboards/covid19/cases

4. Barbier AJ, Jiang AY, Zhang P, Wooster R, Anderson DG. The clinical progress of mRNA vaccines and immunotherapies. Nat Biotechnol. 2022 June 9;40(6):840–54.

5. WHO COVID-19 dashboard [Internet]. [cited 2025 Dec 23]. Available from: https://data.who.int/dashboards/covid19/vaccines

6. Watson OJ, Barnsley G, Toor J, Hogan AB, Winskill P, Ghani AC. Global impact of the first year of COVID-19 vaccination: a mathematical modelling study. Lancet Infect Dis. 2022 Sept 1;22(9):1293–302.

7. World Health Organization. Ten health issues WHO will tackle this year [Internet]. [cited 2026 Jan 10]. Available from: https://www.who.int/news-room/spotlight/ten-threats-to-global-health-in-2019

8. Lazarus JV, Ratzan SC, Palayew A, Gostin LO, Larson HJ, Rabin K, et al. A global survey of potential acceptance of a COVID-19 vaccine. Nat Med. 2021 Feb;27(2):225–8.

9. Lazarus JV, Wyka K, White TM, Picchio CA, Rabin K, Ratzan SC, et al. Revisiting COVID-19 vaccine hesitancy around the world using data from 23 countries in 2021. Nat Commun. 2022 July 1;13(1):3801.

10. Lazarus JV, Wyka K, White TM, Picchio CA, Gostin LO, Larson HJ, et al. A survey of COVID-19 vaccine acceptance across 23 countries in 2022. Nat Med. 2023 Feb;29(2):366–75.

11. Furuse Y, Tabuchi T. Vaccine-related misinformation and attitude toward COVID-19 vaccination in Japan [Internet]. medRxiv. 2024 [cited 2024 July 19]. p. 2024.07.17.24310600. Available from: https://www.medrxiv.org/content/10.1101/2024.07.17.24310600v1.abstract

12. Robinson E, Jones A, Lesser I, Daly M. International estimates of intended uptake and refusal of COVID-19 vaccines: A rapid systematic review and meta-analysis of large nationally representative samples. Vaccine. 2021 Apr 8;39(15):2024–34.

13. Okubo R, Yoshioka T, Ohfuji S, Matsuo T, Tabuchi T. COVID-19 Vaccine Hesitancy and Its Associated Factors in Japan. Vaccines (Basel) [Internet]. 2021 June 17;9(6). Available from: 10.3390/vaccines9060662

14. Schmid P, Böhm R, Das E, Holford D, Korn L, Leask J, et al. Vaccination mandates and their alternatives and complements. Nat Rev Psychol. 2024 Nov 4;3(12):789–803.

15. Volpp KG, Cannuscio CC. Incentives for immunity - strategies for increasing covid-19 vaccine uptake. N Engl J Med. 2021 July 1;385(1):e1.

16. Brewer NT, Buttenheim AM, Clinton CV, Mello MM, Benjamin RM, Callaghan T, et al. Incentives for COVID-19 vaccination. Lancet Reg Health Am. 2022 Apr;8(100205):100205.

17. Khazanov GK, Stewart R, Pieri MF, Huang C, Robertson CT, Schaefer KA, et al. The effectiveness of financial incentives for COVID-19 vaccination: A systematic review. Prev Med. 2023 July;172(107538):107538.

18. World Health Organization. COVID-19 vaccine tracker and landscape [Internet]. [cited 2026 Jan 10]. Available from: https://www.who.int/publications/m/item/draft-landscape-of-covid-19-candidate-vaccines

19. D’Silva C, Fullerton MM, Hu J, Rabin K, Ratzan SC. A global survey to understand general vaccine trust, COVID-19 and influenza vaccine confidence. Front Public Health. 2024 Nov 20;12:1406861.

20. Ino H, Takimoto Y, Nakazawa E. Misinformation targeting replicon vaccine recipients: an urgent public health ethical issue. Lancet. 2024 Nov 16;404(10466):1922–3.

21. Liu C, Tsang TK, Sullivan SG, Cowling BJ, Yang B. Comparative duration of neutralizing responses and protections of COVID-19 vaccination and correlates of protection. Nat Commun. 2025 May 22;16(1):4748.

22. Lazarus JV, White TM, Wyka K, Ratzan SC, Rabin K, Larson HJ, et al. Influence of COVID-19 on trust in routine immunization, health information sources and pandemic preparedness in 23 countries in 2023. Nat Med. 2024 June 29;30(6):1559–63.

23. Ortiz-Prado E, Suárez-Sangucho IA, Vasconez-Gonzalez J, Santillan-Roldán PA, Villavicencio-Gomezjurado M, Salazar-Santoliva C, et al. Pandemic paradox: How the COVID-19 crisis transformed vaccine hesitancy into a two-edged sword. Hum Vaccin Immunother. 2025 Dec 12;21(1):2543167.

24. Leonardelli M, Mele F, Marrone M, Germinario CA, Tafuri S, Moscara L, et al. The effects of the COVID-19 pandemic on vaccination hesitancy: A viewpoint. Vaccines (Basel). 2023 July 3;11(7):1191.

25. Kuppalli K, Omer SB. Measles: the urgent need for global immunisation and preparedness. Lancet. 2025 May 3;405(10489):1565–7.

26. Meda AKR, Medatwal R, Sirohi A, Gupta A, Gupta V, Jain R. Current perspectives on pertussis outbreaks: Causes and consequences. Clin Microbiol Newsl. 2025 Sept 1;52:76–84.

27. Agampodi S, Mogeni OD, Chandler R, Pansuriya M, Kim JH, Excler J-L. Global pandemic preparedness: learning from the COVID-19 vaccine development and distribution. Expert Rev Vaccines. 2024 Jan 30;23(1):761–72.

28. Williams BA, Jones CH, Welch V, True JM. Outlook of pandemic preparedness in a post-COVID-19 world. NPJ Vaccines. 2023 Nov 20;8(1):178.

29. Bollyky TJ, Petersen MB. A practical agenda for incorporating trust into pandemic preparedness and response. Bull World Health Organ. 2024 June 1;102(6):440–7.

30. MacGregor H, Leach M, Desclaux A, Parker M, Grant C, Wilkinson A, et al. Pandemic futures, future preparedness: diverse views in the wake of Covid-19. J Biosoc Sci. 2025 July 24;1–25.

31. 国立感染症研究所. 新型コロナワクチンについて ( 2022 年 3 月 13 日現在 ) [Internet]. [cited 2026 Jan 12]. Available from: https://id-info.jihs.go.jp/niid/ja/2019-ncov-e/11030-covid19-77.html

32. Furuse Y, Tabuchi T. Impact of COVID-19 vaccination by implementation timing and coverage rate in relation to misinformation prevalence in Japan. Vaccine. 2025 May 20;59(127273):127273.

33. Whitaker M, Elliott J, Gerard-Ursin I, Cooke GS, Donnelly CA, Ward H, et al. Profiling vaccine attitudes and subsequent uptake in 1·1 million people in England: a nationwide cohort study. Lancet [Internet]. 2026 Jan 12 [cited 2026 Jan 17];0(0). Available from: 10.1016/S0140-6736(25)01912-9

34. Kaplan RM, Milstein A. Influence of a COVID-19 vaccine’s effectiveness and safety profile on vaccination acceptance. Proc Natl Acad Sci U S A. 2021 Mar 9;118(10):e2021726118.

35. Geers AL, Clemens KS, Colagiuri B, Jason E, Colloca L, Webster R, et al. Do side effects to the primary COVID-19 vaccine reduce intentions for a COVID-19 vaccine booster? Ann Behav Med. 2022 Aug 2;56(8):761–8.

36. Smith LE, Sim J, Sherman SM, Amlôt R, Cutts M, Dasch H, et al. Psychological factors associated with reporting side effects following COVID-19 vaccination: A prospective cohort study (CoVAccS - Wave 3). J Psychosom Res. 2023 Jan 1;164(111104):111104.

37. Ghaznavi C, Yoneoka D, Kawashima T, Eguchi A, Murakami M, Gilmour S, et al. Factors associated with reversals of COVID-19 vaccination willingness: Results from two longitudinal, national surveys in Japan 2021-2022. Lancet Reg Health West Pac. 2022 Oct;27:100540.

38. Seytre B. The difference between COVID-19 vaccine intent and uptake is not associated with exposure to misinformation. Discov Public Health. 2025 Sept 8;22(1):523.

39. Abel ZDV, Roope LSJ, Violato M, Clarke PM. Accuracy of online surveys in predicting COVID-19 uptake and demand: A cohort study investigating vaccine sentiments and switching in 13 countries from 2020 to 2022. Vaccine. 2025 Aug 30;62(127450):127450.

